# Thinking Out Loud: A Qualitative Study of Health Information User Experience in People with Disabilities

**DOI:** 10.64898/2026.03.28.26349601

**Authors:** Sonal S. Sathe, Nathaniel D. Porter, Chreston A. Miller, Michelle S. Rockwell

**Affiliations:** Virginia Tech Graduate School, Blacksburg, VA 24061; University Libraries, Virginia Tech, Blacksburg, VA 24061; Department of Family and Community Medicine, Virginia Tech Carilion School of Medicine, Roanoke, VA 24016

## Abstract

**Background:** People with disabilities use technology, like search engines, to seek health information online. This health information includes information on coronavirus disease, or COVID-19. COVID-19 remains a public health concern. Research shows that people with disabilities encounter frustrations, or “pain points,” when seeking online information, but little is known about these specific pain points and who encounters them.

**Objective:** The goals of this study are to determine pain points for people with disabilities who seek health information online, and to assess how pain points impact the experience of technology use and information seeking.

**Methods:** Ten participants recruited from a prior quantitative survey completed the concurrent think-aloud study over a month-long period. Participants completed four online search tasks and narrated their experiences in real-time while doing so. Transcripts were stored in Taguette; thematic analysis was performed on these transcripts.

**Findings:** Participants were predominantly white, with three identifying as Asian. All ten participants reported having disabilities. Participants with attention deficit hyperactivity disorder (ADHD) reported distracting webpage layout, whereas participants with physical disabilities reported physical fatigue while navigating online information. All participants encountered AI-generated information; only one participant indicated trust in the AI-generated information. Other common sources of information included hospital and governmental webpages, peer-reviewed articles, and news and advertising results. News and advertising results were especially common with respect to search results for “COVID-19 vaccine.” Themes identified included the following: accessibility/usability, AI-generated information, government/hospital and related sources of information, peer-reviewed articles, news and advertising, and sentiment and trust.

**Conclusions:** Information can be fatiguing, distracting, or otherwise difficult to navigate for people with diverse disabilities searching for COVID-19 related information online. Further work should incorporate user feedback from people with disabilities when designing online content.

## Introduction

Technology users with disabilities live in a space where human-computer interaction, health information, and concepts from disability studies, such as universal design, all meet. Understanding users’ needs involves conducting user experience research (UXR). UXR is research “into all possible aspects of an end-user’s experience” of technology [1] and is a component of universal design. Universal design holds a principle of equitable use [2]—that is, the design of environments, including recent technology, should be marketable to people with diverse abilities.[3] When technology and built environments are not designed equitably, people with disabilities face a loss of both access to information and an opportunity to participate in society.[4] A new dynamic of societal exclusion arises when one considers this era to be the age in which artificial intelligence, or AI, is pushed upon consumers by various corporations and entities. UXR should highlight where opportunities exist to introduce and apply the concepts of universal design. In this manner, technology can be equitably designed to benefit society at large.

The integration of AI into digital health information ecosystems has the potential to exacerbate existing disparities in access to and navigation of online health information among people with disabilities. AI enables computers to simulate human behavior and has been embedded into multiple online search engines, like Google and DuckDuckGo, to give conversational-style responses to offer those who use those search engines answers to their questions.[5] AI therefore has already impacted the search habits of technology users.[6] Multiple scholars have shown that AI is biased against people with disabilities [7–9] and people with disabilities encounter difficulties when searching for general health information online.[10–13] People with disabilities may also find the online search results, including AI-generated results, dissatisfying.[14] Additionally, COVID-19 remains more of a threat to people with disabilities than people without disabilities; it is established that people who have disabilities are more likely to experience higher morbidity and mortality from the disease.[15–16] As such, to be a person with a disability seeking health information online about COVID-19 in the age of AI presents a unique set of challenges. When one considers the factors of potentially biased AI, imperfect health information, and the general challenges that people with disabilities face when using technology, one gets a sense of what those challenges might be. However, direct insight to what those challenges are can also come from the people facing those challenges when we ask them what their experiences are. These insights, in turn, can guide more equitable technology design.

An earlier quantitative study [17] provided evidence of people with disabilities finding online content about COVID-19 less satisfying than individuals without disabilities. Surveys, however, provide only a limited window into the experience of technology use and online information among technology users with disabilities. An opportunity exists to dig more deeply into people’s experiences using qualitative methods to collect richer and less mediated data on the lived experience of these technology users. Qualitative research also gives people with disabilities the chance to share their frustrations with using technology in their own voice and words.[18] Some researchers call the frustrations that people encounter with either usability or user satisfaction during their technology use “pain points”.[19] Usability is “the extent to which a product can be used by specified users to achieve specified goals with effectiveness, efficiency, and satisfaction in a specified context of use”.[20] User satisfaction can be thought of as the fulfillment of a user’s specified desire or goal.[21–22]

Qualitative research can be done retrospectively by asking participants to recall events in the past, or in real time, asking participants about the steps they are taking to use the technology in question. Recalling information after an event has passed is less reliable than real-time narration.[23–24] For this study, the goal is to focus on real-time reflections from participants looking for health information about what they are doing and what they may find challenging to navigate when seeking health information online.

Since the availability, accessibility, and usability of health information are part of health equity, people with disabilities must be included in research to achieve public health equity.[25] Unfortunately, this usually does not happen. One systematic review of the use of chatbots for people with disabilities to manage their own wellbeing and overall health further illustrated this point, finding that very few studies considered the wishes of people with disabilities to begin with.[26] Out of 192 studies examined, only 15 included people with disabilities at all.

Qualitative studies must therefore center participant voices to lead the charge in and create positive change towards ensuring that technology is both accessible and usable for people with disabilities.

On the occasions when research does intersect with technology and disability, there is often much left to be desired. One qualitative study of people with cerebral palsy and spinal cord injury focused on the use of mobile health, or mHealth, apps but did not center their voices directly, nor did the researchers themselves indicate any direct relationship to the disability community.[27] Mobile health, or mHealth, is defined as medical and public health practice supported by mobile devices, such as mobile phones, patient monitoring devices, personal digital assistants (PDAs), and other wireless devices.[28]

Previous researchers have qualitatively studied how older adults, who are more likely to have disabilities in general and frequently have multiple disabilities [29–30] interact with data visualizations of COVID-19. However, the rallying cry of the disability rights movement of “nothing about us without us” (along with “our homes, not nursing homes”) also means people with disabilities should be the ones conducting research about disability and that the disability community should have input into the research process in order to shape outcomes that will affect them directly.[31] Yet Fan and colleagues and Zhou and colleagues never indicate they have disabilities themselves or outright give the disability community the voice to shape their desired outcomes. Fan and colleagues and Zhou and colleagues can therefore be said to be doing qualitative user experience research (UXR), but it may not be UXR that allows for the voices and experiences of those in the disability community to take center stage, thereby doing research about the disability community without the disability community’s true input. Also, those prior studies primarily take retrospective analyses as central to their findings, rather than done in real time.

While the studies cited above offer insight into the experience of technology and/or online information in people with disabilities, none of them directly answer whether people with disabilities gain information from AI-embedded tools and use that information to protect themselves from COVID-19 today. We know that COVID-19 remains a risk factor for developing new or further disabilities in both the general population and the disability community. AI is also embedded ever deeper into search engines to generate “overview” results when users seek health information online, and we also know that online health information is not always good or useful. The combination of all three of these may result in frustrating experiences for technology users with disabilities searching for the latest information online on COVID-19 prevention and vaccination.

The goal here, then, was to conduct qualitative UXR that centers the experiences of people with disabilities of searching for online COVID-19 information to find out what frustrations these users encounter while searching for this information. Specifically, the goal of this qualitative UXR study is to determine pain points in the technology/AI-powered space for people with disabilities who are using those assistive tools or ones like them—in real time—to get the information they need. A secondary goal was to assess how those pain points impact the user experience of technology users with disabilities who are searching online for COVID-19 information in hopes of shaping outcomes favorable to the disability community.

Hence, the guiding research questions to answer are:

1. What are the pain points in the user experience of people with disabilities with respect to search engine strategies and results, including AI powered results, to ascertain accurate and usable COVID-19 information?
2. How do those pain points impact the user experience of people with disabilities searching for accurate and usable COVID-19 information online?

Here, accurate COVID-19 information is COVID-19 information that coheres with the information outlined by the World Health Organization about the disease during the public health emergency.[32]

## Methods and Data

### The Concurrent Think-Aloud Method

A think-aloud study invites participants to voice their experiences with a given technology and is ideal for the purposes of giving the participants the chance to speak about their experiences.[33] The think-aloud method is commonly used and widely accepted in qualitative UXR.[33–34] There are two types of think-aloud methodologies: concurrent think-aloud and retrospective think-aloud.[35] In the concurrent think-aloud method, participants verbalize their thoughts about tasks while performing them. In the retrospective method, participants verbalize their thoughts about their tasks after they perform them. The goal of both methods is to capture rich information about the participant’s user experience while letting the participant lead.

There are different strategies people have suggested to ensure optimal information capture.[36] These strategies are adaptable to those who use an augmentative or alternative communication device (AAC) or American Sign Language (ASL) to communicate, so it is not simply a matter of speaking aloud or verbalizing one’s thoughts through a task but in communicating out about the experience. Concurrent think-aloud is used by UX researchers for capturing the user experience in real-time [35] as opposed to the retrospective think-aloud which requires the participant to recall things that have happened in the past.

This study makes space for multiple modalities of communication to allow for a diverse sampling of technology users with disabilities. Communication with technology users will provide insight into their experiences and answer the following questions: Are they able to access information about COVID-19 infection and symptoms, and do they find the information easy to navigate and/or use? How about information on COVID-19 vaccination? More worrisome, what might they be seeing about COVID-19 in relation to the widespread but unsupported belief that ivermectin is an effective treatment or preventative measure for COVID-19?[37] Are they accessing anything that intimates that ivermectin—an anti-parasitic drug—will have any impact on a virus?

To do this, one must allow people with disabilities to narrate freely about what pain points they encounter while using technology to search for COVID-19 information online, which in turn will lead to a broader discussion of the usability experiences with technology in the disability community. As such, data collection proceeded as denoted below.

### Data Collection

Previously, a cross-sectional survey was administered to users of online search engines who sought information about COVID-19. This survey could be taken by adults with and without disabilities; recruitment methods were described by Sathe and Porter.[17] Those who identified themselves as having a disability in that cross-sectional survey were considered as having a disability. If those same respondents indicated that they were interested in participating in a follow-up study, they were recruited to join this concurrent think-aloud study. The disabilities that participants reported could be either developmental or mental health disabilities, like ADHD, autism, or schizophrenia, or could be physical disabilities like type 1 diabetes, multiple sclerosis, postural orthostatic tachycardic syndrome, Crohn’s disease, or similar. Participants who identified themselves as part of a racial or ethnic group, gender identity, and highest education level completed as described in the survey administered by Sathe and Porter [17] were noted as such during data collection for this think-aloud study.

The study took place from April 1 to May 1 of 2025. People without disabilities were not eligible to participate. Other exclusion criteria from the original survey included not being a Virginia resident at the time of the study, being under age 18, and not being able to participate in English.

This study was declared Institutional Review Board (IRB) exempt (Virginia Tech IRB #25-232); the S1 file contains this IRB approval. We opted for verbal consent for our think-aloud study to ensure that we could explain the study to the participants in full and give them the chance to agree or decline accordingly on the day of their session, rather than simply assume they knew what the nature of the study would be. All participants gave informed verbal consent to study participation and to having their participation recorded via Zoom and transcribed. This verbal consent process was approved by the IRB and recorded by the investigator. All participants were informed that any personally identifying information would be kept confidential and would be removed before any deidentified data was shared. Participants were also informed that all study data would be kept for five years after study closure, that there were no foreseeable risks associated with participation, that they could withdraw at any time, and that they would not be penalized for withdrawing if they chose to do so. The study was conducted in accordance with the ethical standards of the Belmont Report and the Declaration of Helsinki.

Participants used their own technologies they would normally use to search, i.e., computers, specialized keyboards, closed captioning, etc; these were open and running for the duration of their participation. Prompts and probes for all participants were predetermined by the research team and are available in the file labeled S2. Working through Zoom, participants either shared their screens as they worked through the questions or read their results aloud from their screen as they worked through the questions. The sessions took about 30 to 45 minutes each to complete.

In the core section of the interview, participants were told to search for four phrases (prompts) in their search engines of choice. In order, those prompts were “COVID-19 infection,” “COVID-19 symptoms,” “COVID-19 ivermectin” and “COVID-19 vaccine”. Participants were asked to verbalize what they were thinking as they conducted each search. No instructions were given regarding whether to avoid personalized search results. Taguette, a free, open-source tool for qualitative research,[38] was used to code and process the participants’ responses. We employed both deductive and inductive codes for the analysis of the participant transcripts.

Pain points were determined by the research team to be frustrations or other issues with usability, accessibility, accuracy, and relevance of the COVID-19 related information that the users found. Pain points, then, were used to guide both deductive and inductive code development. For the deductive codes, the goal was to test the presence of AI-generated information, particularly in relation to information about COVID-19 itself, as well as whether the information was accessible and usable. Therefore, the deductive codes, which were developed before collecting data,[39–40] included “AI-generated,” “information,” “accessibility/usability”, and “COVID”. Inductive codes, developed during coding and analysis, included “advertising,” “news,” and “trust”. Both deductive and inductive codes were discussed and decided by the research team based on the literature cited above; the participant responses also guided the development of inductive codes. These codes helped the researchers show common themes in participant responses.

A thematic analysis was performed as follows: familiarization with the data, initial coding, generating themes, assessing validity and reliability of themes, defining and naming themes, and interpreting and reporting.[41] Reducing biases in this thematic analysis was ensured by peer debriefing throughout the process, but especially for assessing validity and reliability of themes. Revising initial data to verify these themes and eventual conclusions was also employed through the process.

## Data Availability Statement

All ten participant transcripts were submitted to the Qualitative Data Repository (QDR). Participants’ names and other identifying information have been removed to ensure their privacy before submission to the QDR. All participant transcripts are also available in the Supporting Information files (S3-S12). The data deposited in the QDR under DOI: 10.5064/F6CQTZPU will be made public upon publication of this manuscript.

## Results

A total of 10 people participated in this concurrent think-aloud study. One chose to use DuckDuckGo; the rest used Google for their search engines. Further information about the participants can be found in table 1.

**Table 1:** Socio-demographic Characteristics of Participants. Themes generated from coding participant responses are detailed in table 2.

| Socio-demographic Characteristics | Participants (n=10) |
| --- | --- |
| ***Disability Type*** |  |
| Developmental or mental health disability | 5 (50%) |
| Physical disability | 5 (50%) |
| ***Highest Education Level Completed*** |  |
| Did not report/unknown | 2 (20%) |
| No college degree | 1 (10%) |
| Four-year college degree | 2 (20%) |
| Graduate/professional degree | 5 (50%) |
| ***Gender*** |  |
| Female | 7 (70%) |
| Male | 2 (20%) |
| Transgender | 1 (10%) |
| ***Race*** |  |
| White, Caucasian non-Hispanic | 7 (70%) |
| South Asian | 2 (20%) |
| East Asian | 1 (10%) |
| Hispanic | 0 (0%) |
| Black/African-American, non-Hispanic | 0 (0%) |
| Native Hawaiian/Pacific Islander or<br>American Indian | 0 (0%) |
| *Age Range* |  |
| Did not report/unknown | 2 (20%) |
| 18-24 | 2 (20%) |
| 25-34 | 5 (50%) |
| 35-44 | 1 (10%) |
| 45 and up | 0 (0%) |

**Table 2:** Themes and Subthemes/Pain Points from Participant Responses. Each theme is discussed in more detail below, along with representative quotes from participants.

| **Themes** | **Subthemes/Pain Points** |
| --- | --- |
| 1. Accessibility/usability of online<br>information | Pain points: inability to use<br>assistive technology effectively, |
|  | distracting layout, or information too tiring or too much to navigate |
| 2. AI-Generated Information | Subtheme: sourcing of information |
| 3. Hospital, Government, and Related Sources | Subtheme: sourcing and date of information |
| 4. Peer-Reviewed Articles | Subtheme: currency of article |
| 5. News and Advertising | Subtheme: local, national, and international news sources<br>Pain points: irrelevancy/outdated information |
| 6. Sentiment and Trust | Subtheme: trust and mistrust of online information |

### Accessibility and Usability of Online Information

#### Theme 1: Accessibility and Usability of Information

All participants indicated that the information they searched for and found was accessible to them with any assistive devices they had in use. However, participants reported issues with usability of online information, especially upon completion of searching “COVID-19 ivermectin” and “COVID-19 vaccine”, respectively. As far as specific usability issues, distracting layouts of the pages linked from the results were more likely to be mentioned by participants who reported having ADHD (n=5). People with physical disabilities were likely to report fatigue while searching for information (n=3).

With respect to both a distracting layout and a wider accessibility and usability issue at large, one participant with ADHD commented:

> *“Yeah, I would have had to use the [screen] reader. And I know for a fact, like some websites, like I can’t even use the reader, like it won’t let me use my reader. So, I have to download it. Or like print it as a PDF. And then do that. And still, it’s like very well… It drives me crazy because I’m reading a big text…”*

Another participant with ADHD shared their experience of the information they found from all four prompts, but especially the last one which was “COVID-19 vaccine”:

> *“Yeah, it was all kind of jumbled up and everything. Like, it wasn’t very, it could have been more organized. Or something like that… Cleaner. Where… I’m not searching everywhere for stuff.”*

Another participant with ADHD noted that the information they received from all four search prompts was difficult to process:

> *“I think I… if I was in a pinch and I needed to… learn something really quick about COVID, then I could use the information, but it would take me a lot longer to understand, like gain a deeper understanding.”*

Concerns about physical fatigue during the search tasks were reported by participants who reported physical disabilities, like type 1 diabetes (n=2) or postural orthostatic tachycardia syndrome, or POTS for short (n=1). One participant with type 1 diabetes stated:

> *“I can usually stay focused for about an hour. Any longer than that is like rough territory My disability is, I mean, I’m type one diabetic and so it’s not predictable when there’s going to be a problem and what the problem is going to be…”*

All the above results indicate that while COVID-19 related information is available, technology users with disabilities may have trouble processing these results due to concerns with the way the information arises while searching online, especially if they have ADHD. In other words, these technology users may find results accessible but not usable or useful. They may also face physical difficulties if their search results take a long time to go through, or simply not be willing to take the time to go through the results in the first place. Issues with concentration, organization, and memory are common among many disability types, including those with ADHD, brain injuries, and disabilities that cause fatigue or brain fog.[42]

### Information Sources

Participants felt that the information they found with different prompted searches may be difficult to process, may or may not come from trusted sources, and might simply be too much to process or tiresome to navigate. Specifically, the results indicate that the pain points in the user experience of online search information in technology users with disabilities include AI-generated results, distracting webpage layout, user fatigue, and the nature and sourcing of the information they received. These results have implications for future user experience work with the disability community.

#### Theme 2: AI-Generated Information

All participants encountered AI-generated information in their very first result for their first search. AI-generated information was also the first result for all Google search engine users for the second prompt, “COVID-19 symptoms”.

The sole DuckDuckGo user received AI-generated results as an option for the results from the third prompt as well, although DuckDuckGo did offer an “off” option if the user did not want to see an AI-generated result. They even noted:

> *“[The] first thing is an AI-assisted answer, but it is at least asking me if I want to do that rather than doing it on purpose automatically.”*

For the results from the first prompt for all users, the source which the AI was pulling from was also not stated 50% of the time. For the results from the second prompt, sources varied but were stated in the AI result—even if they also came up later in the search results page. One participant even noted:

> *“My first, so my first thing again is AI. But interestingly um the one that shows up immediately after that has… the exact same information like bullet point by bullet point that AI is giving me…mixture of sources.”*

In all cases, it was clear to the participants that the AI-generated results were AI-generated, but the statement indicating that a given paragraph of text was created using AI was at the bottom of said AI-generated results in very small print for both prompts 1 and 2.

The DuckDuckGo user did not state where the AI label was in their search results for prompts 1, 2, and 3, though they did state that the AI-generated result for prompt 2 drew from at least two sources, one of which was Wikipedia. None of the participants encountered an AI-generated result for the final prompt, which was “COVID-19 vaccine”.

Issues with user fatigue were still the case with AI-generated results just as with non-AI generated results. However, the information overload for people already fatigued with AI factored into the equation may predispose a tired or distracted online search engine user to opt for unverified information to make health decisions.[42]

#### Theme 3: Hospital, Government, and Related Sources

Both hospital and government webpages came up directly for all participants in response to all four prompts, though these results were far more likely to result from the first three prompts than the last. Results from the Food and Drug Administration (FDA) popped up for all Google users for the first result from the third prompt, clearly stating that ivermectin was not approved by the FDA for treating COVID-19. Results from the Centers for Disease Control and Prevention (CDC) typically arose in relation to prompts 1, 2, and sometimes prompt 3 and 4 as well, depending on the participant. Mayo Clinic results routinely came up for results from prompts 1 and 2, and in at least four cases, University of California at Los Angeles Health was a result for searches arising from prompt 3. The National Institutes of Health (NIH) webpage tended to result from users searching with prompts 3 and 4.

All participants noted the date of publication of each of these webpages if present. The FDA webpage was published in 2024, whereas some of the other webpages dated from 2021 or 2020. While nothing explicitly came up about the validity of the information based on how old the information was, participants noted they expected certain results. One participant commented:

> *“And then following that, my third one is from NIH. Which is another government website…I feel like it’s very standard for what you would see with anything entered for COVID.”*

Some government-related webpages arose as part of the results from prompt 3, and not all users were able to discern where they were from. One webpage in which this routinely happened was the National Health Service (NHS) from the United Kingdom; at least three participants did not know what this webpage was or what the abbreviation stood for.

For the DuckDuckGo user, results from using prompts 1 and 2 did yield Mayo Clinic [43] and CDC [44] webpages, but for prompt 1 they also received some information from the World Health Organization, dated 2023 [45]; the World Health Organization is an inter-governmental organization.

All participants, when they encountered a hospital or government-related webpage, had the opportunity to click on links to find out more information. These links took the participants directly to webpages where they were able to find what they were looking for.

#### Theme 4: Peer Reviewed Articles

Peer reviewed articles most frequently arose in the results from “COVID-19 ivermectin”, especially for all Google users. Some were directly displayed in the search results; others were linked through Google Scholar. All participants were able to access an open-access article published in the *New England Journal of Medicine*, though many articles in that journal are not open-access, and therefore not readily available to the broader community. Other journals were not mentioned by name, but three other participants noted the existence of a systematic review in one of these articles that came up in the search results.

For the articles arising from “COVID-19 ivermectin”, most participants did note that the research article in the *New England Journal of Medicine* was from 2020, but only upon the researcher probing the participants for further information about the article. One participant also struggled with examining the nature of a published article, which turned out to be an editorial:

> *“It didn’t say systematic review in the paper. In the title. But since it doesn’t have any like original data, so I thought it would be a review paper”*

Another participant examined an article beyond the title, but noted:

> *“Yeah, there’s like a label for it [research article]. But it’s the same label as the introduction and the methods and stuff.”*

One other participant who had multiple physical disabilities noted that they found articles that had more to do with immunotherapeutic studies and pathology; they did not specify the publication date of those articles.

All told, about half the articles the participants found were published after the public health emergency was ended by the World Health Organization in 2023, and participants did not always take the time to read these articles beyond the title.

#### Theme 5: News and Advertising

News results and sponsored advertising results arose for all participants upon finding search results for “COVID-19 vaccine”, though one participant got news results from “COVID-19 symptoms” as well. One participant reported a news source from News India Times, which is a weekly newspaper focused on news from and about India; the participant was not of Indian origin. Other participants (n=3) reported Reuters as a news source regarding the latest on Novavax and Pfizer vaccines as well as the latest news on efforts to impact the release of the Novavax and Pfizer vaccines, with one participant commenting:

> *“..one of the resources from two days ago…says RFK Jr. eyes reversing CDC’s COVID vaccine recommendation for children…”*

Some participants (n=2) stated that Fox News affiliates ended up in their search results as well. Not all news-related results were well received. One participant noted:

> *“I wish that the first results weren’t news articles a lot of the time… I think that it’s easy for people to see that and take dramatic current news as the guidelines they should be going by rather than actual verified information.”*

Another participant encountered a news affiliate that was not in the geographic area of study, or any place they had stated they had lived in the past, in their search results. They commented:

> *“Fox Nashville…I’m not someone who typically uses that news source so that’s weird.”*

As such, a mix of news sources arose upon these technology users seeking information about the latest COVID-19 vaccine, and user reactions to these news sources varied. Sponsored advertising results for the latest COVID-19 vaccine—at the time of this study, the latest COVID-19 vaccines were the Pfizer version—came up for all Google users; the DuckDuckGo user did not report seeing advertisements in any of their results. Of the other nine participants searching with Google, three participants stated that pharmacies advertising the latest COVID-19 vaccine, and its availability, showed up in the first three results after searching “COVID-19 vaccine”. The most common pharmacies advertising the COVID-19 vaccine were Kroger, Walgreens, and CVS; the most common vaccine types advertised were Pfizer and Moderna. One participant stated their immediate thoughts regarding seeing some of the results:

> *“Oh, we’re selling this. Come here. Please come to us. I don’t know. I know it’s still promoting the vaccine, which is fine, but I still feel like it, I don’t know… I thought I’d get a little bit more of, you know, what we were kind of alluding to more concrete …more factual based.”*

While the vaccine information through advertisements and news was welcome by all the participants, the news and advertising surrounding the vaccine was irrelevant to participants who were searching for information about the latest vaccine itself, rather than the locations they could go to get the COVID-19 vaccine. Not all locations that provide the COVID-19 vaccine are accessible to people with disabilities, either, which the search results did not account for.

### Emotional Reactions

#### Theme 6: Sentiment and Trust

Participants expressed negative sentiments around known misinformation relating to COVID-19 prevention and control, Fox News, and/or anti-vaccine propaganda. Some anti-vaccine propaganda includes the hoax that vaccines cause autism.[46] Considering the gross misinformation surrounding vaccines and autism, one participant, upon beginning the search for the prompt “COVID-19 vaccine”, stated:

> *“Okay. Covid-19 vaccine. If something comes up with autism, I’m just going to lose it.”*

Similar sentiments were expressed by two other participants when searching for the third prompt, “COVID-19 ivermectin”, since ivermectin is not supported by scientific evidence as treatment for COVID-19. [47] One participant stated:

> *“One of my friends … kind of believes all this …my other friend…kind of sat down with him and then he explained…. But he still believes that it is all conspiratory and that ivermectin is the cure to healing the world… but parasites … different construction than viruses.”*

Regarding misinformation about COVID-19 from news sources, another participant stated:

> *“I don’t think I would trust it. I would read it for like enjoyment rather than information.”*

As far as trust in the actual information sources, participants reported a high level of trust in government and hospital websites (n=6), with one participant noting when asked where the information about the vaccine was coming from:

> *“I’d say trusted sites. The CDC, WHO, and Mayo Clinic.”*

One participant, in contrast to the other nine participants, expressed trust in AI-generated results outright:

> *“I think the top thing that I can think about as it relates to Google searches is the AI responses. And I think that’s something that I tend to have trust in.…That the AI overview, especially when it comes to questions like the COVID-19 symptoms or you know, like just generally COVID-19 infection like what is that I tend to trust the AI overview.”*

Trust in various sources varied among participants, but most of them regarded information about ivermectin treating COVID-19 as false/misinformation. The biggest problem participants faced was finding up-to-date information and vaccine information that matched what they were seeking.

In sum, across the six themes discussed above, issues were present as follows: first, user distraction and fatigue; second, AI-generated information that did not always have a date attached; and third, information quality and sourcing. Taken together, these pain points make for a suboptimal user experience for the technology user with a disability who is seeking information about COVID-19 online.

## Discussion

### General

Based on the findings from this concurrent think-aloud study, user experience work would benefit from co-designing technology with neurodivergent users to make the layout of pages less distracting. There is an opportunity for user fatigue in technology users with disabilities to be accounted for when thinking of search-related strategies to find information through co-design with these users as well, since user interfaces—the point at which humans interact with technology [48] — can be redesigned with people with disabilities in mind. Doing so would make technology users less overwhelmed when viewing health information online; in turn, they may be able to digest health information more easily and therefore make health decisions more easily. The volume of the information received and the fatigue in navigating information also impacted user willingness to persist in engaging in the digital space until they found results from the search prompts detailed above.

Participants in this study were not always reading scientific literature critically. The first two participants especially struggled to assess the nature of some of the peer-reviewed literature they found online, and one of those two is in a doctoral program. Further work should not only focus on teaching the public–and graduate students— to critically review and examine the results of peer-reviewed studies, but also in making the information from peer-reviewed research easier to both access and understand to both graduate students and the public. Efforts in both the areas of making information accessible and comprehensible, and efforts in teaching graduate students to read critically for information, feed into existing endeavors, like open science,[49] to increase overall trust in science and medicine for both the public and researchers. Further work could also dive more specifically into the relationship that technology users with disabilities have with AI-powered tools and searches in general.

In short, finding useful, accurate and timely health information online relies on a locus of factors only partly in the seeker’s control. Some of these challenges are further exacerbated by disabilities, which reduces both functional accessibility and usability in ways that include not only traditional digital accessibility concerns like not being accessible for screen reader users, but also often-ignored issues like ease of information organization and risk of fatigue. We can infer that the interaction of unusability of online content, levels of trust in information sources, and the information sources themselves make seeking and comprehending COVID-19 related information difficult for technology users with disabilities. Such difficulties may contribute to delayed or no vaccination against COVID-19. Without vaccination, people with disabilities remain at elevated risk from COVID-19 related illness and death. As such, efforts to make health information easy to digest would likely increase vaccination rates and thereby ameliorate a situation that would exacerbate a public health crisis.

## Limitations and Future Opportunities

Future work should also consider keeping the concurrent think-aloud methodology as a starting gate to a larger qualitative study. A focus group of users with the same disability would be ideal for such an endeavor. These focus groups should include users with vision, hearing, and motor disabilities, since these users were underrepresented in this study. Further work should also expand geographic reach beyond Virginia.

Additionally, most participants who reported mental health disabilities in the prior quantitative study did not participate in this concurrent think-aloud study. Only one participant in this think-aloud study reported a mental health diagnosis. Stigma may be a barrier for those who have mental health disabilities to participate in research,[50] even though lived experience research with people with mental health disabilities has become a research area focus for some scholars.[51]

Further work should investigate ways to overcome barriers to research engagement in people with mental health disabilities. One way to do this is to focus on the application of dissemination and implementation science.[52] Dissemination is the active approach of spreading evidence-based interventions to the target audience via determined channels using planned strategies; implementation is focused on strategies and factors that support the initial integration and use of research findings into practice within specific real-world contexts.[53] Dissemination and implementation science therefore pairs both dissemination and implementation to improve healthcare.[53] To make sure that people can even see providers in the first place, public health and healthcare researchers should either begin or continue efforts to disseminate and implement high value healthcare, and de-implement low-value healthcare, by determining and eliminating existing barriers to healthcare provider access.[54] The elimination of unnecessary policies and practices that make it difficult to access healthcare services, called sludge,[55] is also a crucial part of eliminating existing barriers to accessing healthcare. Some researchers have begun the process of examining existing sludge in the healthcare system with respect to colon cancer screenings.[56] Follow-ups of these audits should be conducted, especially to reduce waiting times to see physicians. Similar audits and changes based on these audits should be conducted in other areas of primary care.

With only ten participants, this study cannot provide a full picture of the diversity of user experience of technology in the entire disability community, but it does provide new data. Further research should aim for studies that involve interviews, ethnographic methods, grounded theory, or perhaps even community-based participatory research (CBPR) for a deeper insight into the challenges that these technology users face when using technology to search for health information online. Further studies focused on specific subpopulations of the disability community could also be valuable, both because they could gather more data on the experience of one group and because they can ensure that group is more comparable and well-represented than they are in this study.

Finally, it is important to consider that nine out of these ten participants had completed at least a four-year college degree. Results may be quite different from a sample where most participants have completed less education than a college degree. It is also worth noting here that in the disability community, college degree holders are a small minority; students with disabilities encounter significant barriers to degree completion.[57] Also, only three out of ten participants were nonwhite (one East Asian, two South Asian). Results would look quite different in a more racially and ethnically diverse sample. Further work should aim to recruit more nonwhite participants in research. Perhaps building community partnerships with black disability, indigenous disability, or other communities may help to build trust and increase research participation rates in those communities.[58]

## Conclusions

To sum up, technology users with disabilities encounter a wide variety of sources, including AI-generated information, when they search for COVID-19 related information. Other sources of information that technology users with disabilities encounter include hospital/government and related sources, peer-reviewed articles, and news and sponsored advertising results. Not all this information is easily navigable for people with disabilities, who might feel fatigued or distracted upon encountering search results. Additionally, not all the information in the digital space is up to date; the lack of current information could hamper information gathering for all users. Future work should incorporate user feedback to make information accessible, usable, and satisfying for technology users with disabilities so they can access COVID-19 information to protect themselves against the disease.

## Data Availability

All relevant data are within the manuscript as well as the Supporting Information files.

https://doi.org/10.5064/F6CQTZPU

## Acknowledgments

Special thanks to Ashley Shew of the Department of Science, Technology, and Society at Virginia Tech for editing for length and clarity.

## References

1. Kaplan K. What Is User Experience (and What Is It Not)? [Internet]. Nielsen Norman Group. 2024. Available from: https://www.nngroup.com/articles/what-is-user-experience/

2. Mace R, Hardie G, Place J. Toward universal design. Oxfordshire, UK: Taylor & Francis Ltd.; 1991.

3. Vouchilas G. Contemporary Living: Bringing a Universal Design Principle into the Mix. Journal of Family and Consumer Sciences [Internet]. 2017 [cited 2026 Mar 10];109(4):21–5. doi: 10.14307/JFCS109.4.21

4. Nielsen AF, Landa-Mata I. Expanding the understanding of universal design beyond technical solutions and physical environment – 8 policy intervention areas. Transport Policy. 2025 Jun; 167:157–77. doi: 10.1016/j.tranpol.2025.03.028

5. Nelson HC, Beauchamp MT, Pace AA. The Reliability Gap: How Traditional Search Engines Outperform Artificial Intelligence (AI) Chatbots in Rosacea Public Health Information Quality. Cureus [Internet]. 2025 Jun 22. doi: 10.7759/cureus.86543

6. Brown J, Moran K, Rosala M. How AI Is Changing Search Behaviors [Internet]. Nielsen Norman Group. 2025. Available from: https://www.nngroup.com/articles/ai-changing-search-behaviors/

7. Packin NG. Disability Discrimination Using AI Systems, Social Media and Digital Platforms: Can We Disable Digital Bias? SSRN Electronic Journal. 2020. doi:10.2139/ssrn.3724556

8. Manzoor R, Hussain W, Anjum ML. Out of dataset, out of algorithm, out of mind: a critical evaluation of AI bias against disabled people. AI & Society. 2024 Dec 28. doi:10.1007/s00146-024-02168-8

9. Williams DP. Disabling AI: Biases and Values Embedded in Artificial Intelligence. In: Gunkel DJ, editor. Handbook on the Ethics of Artificial Intelligence [Internet]. Edward Elgar Publishing; 2024 [cited 2026 Feb 27]. p. 246–61. doi:10.4337/9781803926728.00022

10. Kim H, Hwang H, Sojung Gwak, Yoon J, Park K. Improving communication and promoting social inclusion for hearing-impaired users: Usability evaluation and design recommendations for assistive mobile applications. PloS one. 2024 Jul 17;19(7):e0305726–6. doi: 10.1371/journal.pone.0305726

11. Mulliken A. “There is Nothing Inherently Mysterious about Assistive Technology”: A Qualitative Study about Blind User Experiences in US Academic Libraries. Reference & User Services Quarterly. 2017 Dec 28;57(2):115.

12. Spiel K, Hornecker E, Williams RM, Good J. ADHD and technology research–investigated by neurodivergent readers. In Proceedings of the 2022 CHI conference on human factors in computing systems 2022 Apr 29 (pp. 1–21). doi:10.1145/3491102.3517592

13. Ortiz-Escobar LM, Chavarria MA, Schönenberger K, Hurst S, Stein MA, Mugeere A, et al. Assessing the implementation of user-centred design standards on assistive technology for persons with visual impairments: a systematic review. Front Rehabil Sci. 2023 Sep 6;4:1238158. doi:10.3389/fresc.2023.1238158

14. Mergen A, Çetin-Kılıç N, Özbilgin MF. Artificial Intelligence and Bias Towards Marginalised Groups: Theoretical Roots and Challenges. In: Vassilopoulou J, Kyriakidou O, editors. International Perspectives on Equality, Diversity and Inclusion [Internet]. Emerald Publishing Limited; 2025 [cited 2026 Feb 27]. p. 17–38. doi:10.1108/S2051-233320250000012004

15. Kamalakannan S, Bhattacharjya S, Bogdanova Y, Papadimitriou C, Arango-Lasprilla J, Bentley J, et al. Health Risks and Consequences of a COVID-19 Infection for People with Disabilities: Scoping Review and Descriptive Thematic Analysis. IJERPH. 2021 Apr 20;18(8):4348. doi:10.3390/ijerph18084348

16. Shakespeare T, Ndagire F, Seketi QE. Triple jeopardy: Disabled People and the COVID-19 Pandemic. The Lancet [Internet]. 2021 Apr;397(10282):1331–3. doi: 10.1016/S0140-6736(21)00625-5

17. Sathe S, Porter N. By the Numbers: Results from A Quantitative Study of Health Information User Experience in People with and Without Disabilities [Internet]. Public and Global Health; 2025 [cited 2026 Feb 27]. doi:10.1101/2025.10.01.25337080

18. Ashby CE. Whose “Voice” is it Anyway?: Giving Voice and Qualitative Research Involving Individuals that Type to Communicate. DSQ. 2011 Oct 25;31(4). doi:10.18061/dsq.v31i4.1723

19. Gibbons S. Three Levels of Pain Points in Customer Experience [Internet]. Nielsen Norman Group. 2021. Available from: https://www.nngroup.com/articles/pain-points/

20. Grassi PA, Garcia ME, Fenton JL. Digital identity guidelines: revision 3 [Internet]. Gaithersburg, MD: National Institute of Standards and Technology; 2017 Jun [cited 2026 Feb 27]. p. NIST SP 800-63-3. Report No.: NIST SP 800-63-3. Available from: https://nvlpubs.nist.gov/nistpubs/SpecialPublications/NIST.SP.800-63-3.pdf doi:10.6028/NIST.SP.800-63-3

21. Kelly D. Methods for evaluating interactive information retrieval systems with users. Foundations and Trends® in Information Retrieval. 2009 Apr 28;3(1–2):1–224.

22. Siro C, Aliannejadi M, Maarten de Rijke. Understanding and Predicting User Satisfaction with Conversational Recommender Systems. ACM Transactions on Information Systems. 2023 Nov 8;42(2):1–37. doi: 10.1145/3624989

23. Marsden PV, editor. Handbook of survey research. 2. ed. Bingley: Emerald Group Publ; 2010. 886 p.

24. Howard RW. Testing the accuracy of the retrospective recall method used in expertise research. Behav Res. 2011 Dec;43(4):931–41. doi:10.3758/s13428-011-0120-x

25. Havercamp SM, Krahn GL, Murray AJ, Akobirshoev I, Bellamy CD, Bonardi A, et al. A call to action to include disability in intersectional health equity research and policy. The Lancet Regional Health - Americas. 2025 Sep;49:101199. doi:10.1016/j.lana.2025.101199

26. Federici S, De Filippis ML, Mele ML, Borsci S, Bracalenti M, Gaudino G, et al. Inside pandora’s box: a systematic review of the assessment of the perceived quality of chatbots for people with disabilities or special needs. Disability and Rehabilitation: Assistive Technology. 2020 Oct 2;15(7):832–7. doi:10.1080/17483107.2020.1775313

27. Zhou L, Saptono A, Setiawan IMA, Parmanto B. Making Self-Management Mobile Health Apps Accessible to People With Disabilities: Qualitative Single-Subject Study. JMIR Mhealth Uhealth. 2020 Jan 3;8(1):e15060. doi:10.2196/15060

28. World Health Organization. WHO guideline Recommendations on Digital Interventions for Health System Strengthening [Internet]. PubMed. Geneva: World Health Organization; 2019. Available from: https://www.ncbi.nlm.nih.gov/books/NBK541902/

29. Fan M, Wang Y, Xie Y, Li FM, Chen C. Understanding How Older Adults Comprehend COVID-19 Interactive Visualizations via Think-Aloud Protocol. International Journal of Human–Computer Interaction. 2023 May 9;39(8):1626–42. doi:10.1080/10447318.2022.2064609

30. Brucker DL, Lauer E, Boege S. Americans Aging with Disabilities Are More Likely to Have Multiple Chronic Conditions. Journal of Disability Policy Studies. 2023 Jun;34(1):52–60. doi:10.1177/10442073221107079

31. Hofmann M, Kasnitz D, Mankoff J, Bennett CL. Living Disability Theory: Reflections on Access, Research, and Design. In: Proceedings of the 22nd International ACM SIGACCESS Conference on Computers and Accessibility [Internet]. Virtual Event Greece: ACM; 2020 [cited 2026 Feb 27]. p. 1–13. doi:10.1145/3373625.3416996

32. World Health Organization. Coronavirus [Internet]. www.who.int. 2025. Available from: https://www.who.int/health-topics/coronavirus#tab=tab

33. Nielsen J. Thinking Aloud: The #1 Usability Tool [Internet]. Nielsen Norman Group. 2012. Available from: https://www.nngroup.com/articles/thinking-aloud-the-1-usability-tool/

34. Fan M, Tibdewal V, Zhao Q, Cao L, Peng C, Shu R, et al. Older Adults’ Concurrent and Retrospective Think-Aloud Verbalizations for Identifying User Experience Problems of VR Games. Interacting with Computers. 2022 Feb 1;34(4):99–115. doi:10.1093/iwc/iwac039

35. Fan M, Lin J, Chung C, Truong KN. Concurrent Think-Aloud Verbalizations and Usability Problems. ACM Trans Comput-Hum Interact. 2019 Oct 31;26(5):1–35. doi:10.1145/3325281

36. Noushad B, Van Gerven PWM, De Bruin ABH. Twelve tips for applying the think-aloud method to capture cognitive processes. Medical Teacher. 2024 Jul 2;46(7):892–7. doi:10.1080/0142159X.2023.2289847

37. Bibbins-Domingo K, Malani PN. At a Higher Dose and Longer Duration, Ivermectin Still Not Effective Against COVID-19. JAMA. 2023 Mar 21;329(11):897. doi:10.1001/jama.2023.1922

38. Rampin R, Rampin V. Taguette: open-source qualitative data analysis. JOSS. 2021 Dec 10;6(68):3522. doi:10.21105/joss.03522

39. Bingham A. Deductive and Inductive Approaches to Qualitative Data Analysis: The Five-Cycle Process. In: Proceedings of the 2021 AERA Annual Meeting [Internet]. AERA; 2021 [cited 2026 Feb 27]. doi:10.3102/1682697

40. Bingham AJ. From Data Management to Actionable Findings: A Five-Phase Process of Qualitative Data Analysis. International Journal of Qualitative Methods. 2023 Oct;22:16094069231183620. doi:10.1177/16094069231183620

41. Cernasev A, Axon DR. Thematic Analysis in Qualitative Research: An overview. Research and scholarly methods: Thematic analysis. 2023 May 12;6(7):751–5. doi:10.1002/jac5.1817

42. Callan C, Ladds E, Husain L, Pattinson K, Greenhalgh T. ‘I can’t cope with multiple inputs’: a qualitative study of the lived experience of ‘brain fog’ after COVID-19. BMJ open. 2022 Feb 1;12(2):e056366. doi: 10.1136/bmjopen-2021-056366.

43. Mayo Clinic. Coronavirus Disease 2019 (COVID-19) [Internet]. Mayo Clinic. 2024. Available from: https://www.mayoclinic.org/diseases-conditions/coronavirus/symptoms-causes/syc-20479963

44. Centers for Disease Control and Prevention. Symptoms of COVID-19 [Internet]. COVID-19. CDC; 2024. Available from: https://www.cdc.gov/covid/signs-symptoms/index.html

45. World Health Organization. Coronavirus Disease (COVID-19) [Internet]. World Health Organization. 2023. Available from: https://www.who.int/news-room/fact-sheets/detail/coronavirus-disease-(covid-19)

46. Gabis LV, Attia OL, Goldman M, Barak N, Tefera P, Shefer S, et al. The myth of vaccination and autism spectrum. European Journal of Paediatric Neurology. 2022 Jan;36:151–8. doi:10.1016/j.ejpn.2021.12.011

47. Perlis RH, Lunz Trujillo K, Green J, Safarpour A, Druckman JN, Santillana M, et al. Misinformation, Trust, and Use of Ivermectin and Hydroxychloroquine for COVID-19. JAMA Health Forum [Internet]. 2023 Sep 29;4(9):e233257. doi: 10.1001/jamahealthform.2023.3257

48. Sutcliffe A. Designing for User Engagement. Springer Nature; 2022.

49. Song H, Markowitz DM, Taylor SH. Trusting on the shoulders of open giants?Open science increases trust in science for the public and academics. Journal of Communication. 2022 Jun 30;72(4):497–510. doi: 10.25417/uic.19750294

50. Baxter L, Burton A, Fancourt D. Community and cultural engagement for people with lived experience of mental health conditions: what are the barriers and enablers? BMC Psychol. 2022 Dec;10(1):71. doi:10.1186/s40359-022-00775-y

51. Davis K, Balasuriya L, Dixon LB. Engaging People With Lived Experience in Mental Health Services and Research. PS. 2022 Apr 1;73(4):476–7. doi:10.1176/appi.ps.22073001

52. Shelton RC, Lee M, Brotzman LE, Wolfenden L, Nathan N, Wainberg ML. What Is Dissemination and Implementation Science?: An Introduction and Opportunities to Advance Behavioral Medicine and Public Health Globally. International Journal of Behavioral Medicine. 2020 Feb;27(1):3–20. doi: 10.1007/s12529-020-09848-x.

53. Durlak JA, DuPre EP. Implementation Matters: A Review of Research on the Influence of Implementation on Program Outcomes and the Factors Affecting Implementation. American Journal of Community Psychology. 2008 Mar 6;41(3-4):327–50. doi: 10.1007/s10464-008-9165-0.

54. Shato T, Kepper MM, McLoughlin GM, Tabak RG, Glasgow RE, Brownson RC. Designing for Dissemination among Public Health and Clinical Practitioners in the United States. Journal of clinical and translational science [Internet]. 2023 Dec 14;8(1):1–26. doi:10.1017%2Fcts.2023.695

55. Sunstein, C.R. Sludge Audits. Behavioural Public Policy. 2022;6(4):654–73. doi:10.1017/bpp.2019.32

56. Rockwell MS, Frazier MC, Stein JS, Dulaney KA, Parker SH, Davis GC, Rockwell JA, Castleman BL, Sunstein CR, Epling JW. A ‘Sludge Audit’for health system colorectal cancer screening services. The American Journal of Managed Care. 2023 Jul 1;29(7):e222. doi: 10.37765/ajmc.2023.89402

57. Carroll JM, Pattison E, Muller C, Sutton A. Barriers to Bachelor’s Degree Completion among College Students with a Disability. Sociological Perspectives. 2020 Mar 5;63(5):809–32. doi: 10.1177/0731121420908896

58. Tamlyn AL, Tjilos M, Bosch NA, Barnett KG, Perkins RB, Walkey A, et al. At the intersection of trust and mistrust: A qualitative analysis of motivators and barriers to research participation at a safety-net hospital. Health Expectations. 2023 Jun;26(3):1118–26. doi:10.1111/hex.13726

